# Phasic containment of COVID-19 in substantially affected states of India

**DOI:** 10.1101/2020.05.05.20092130

**Authors:** Manisha Mandal, Shyamapada Mandal

## Abstract

The spread of COVID-19 epidemic in some highly-impacted Indian states displayed a characteristic sub-exponential growth projected up to 3 May 2020, as a consequence of lockdown strategies, in addition to improvement of reproduction number (R), serial interval, and daily growth rate, but not case fatality rate (CFR). The effect of COVID-19 containment was more prominent in second phase of lockdown with declining R, which was still >1, suggesting the requirement of sustained interventions for effective containment of COVID-19 pandemic in Indian context.

## Introduction

Since the first detection of COVID-19 case on 30 January 2020, India reported 26,917 confirmed cases and 826 deaths; Maharashtra (7628) remains the worst affected state, followed by Gujarat (3071), Delhi (2625), and Rajasthan (2083), as of 26 April 2020.^1^ India, in an attempt to manage this crisis, enacted a two-phase nationwide lockdowns, from 25 March 2020 to 14 April 2020, and thereafter extended up to 3 May 2020, because of the escalating spread of COVID-19 in India, in 32 states/union territories.^2,3^ In view of this, we represent the epidemic course of the disease in some highly-impacted Indian states, and its key features, using real data and model-based prediction, and explore the effectiveness of lockdowns in India on COVID-19 pandemic, since no such studies are available in India.

## Methods

Data on COVID-19, in Indian states, including Maharashtra, Delhi, Gujarat, Madhya Pradesh, Tamil Nadu, Rajasthan, and Uttar Pradesh (with >1000 confirmed cases, as of April 20, 2020), were retrieved electronically from publicly available domain of the Ministry of Health and Family Welfare, Government of India,^1^ from 30 January 2020 through 20 April 2020^1^; Kerala and West Bengal having case numbers <1000 were also included in the study. Daily growth rate (DGR), case fatality rate (CFR), serial interval (SI), and time-varying reproduction number (R) of COVID-19 cases were estimated, before and after lockdown, following the criteria mentioned earlier.^4^

## Results

The spread of COVID-19 displayed a characteristic sub-exponential linear growth (mean DGR: 0.06; range: 0.01, for Kerala, to 0.11, for Gujarat, 95% CI: 0.04-0.08, 11 April 2020 to 3 May 2020) that deviates from exponential growth estimates (mean DGR: 0.1643; range: 0.1163, for Kerala, to 0.2175, for Tamil Nadu, 95% CI: 0.1392-0.1894, 2 March 2020 to 3 May 2020), as a consequence of lockdown strategies (Figure 1). Reduction of R, from a range of 1.35-2.86 (pre-lockdown 95% CI: 1.49-2.21) to 1.13 - 1.67, in first phase, and to 1.08-1.63, in second phase of lockdowns (post-lockdown 95% CI: 1.20-1.42), and cases by 56%-98%, in individual states, have been reflected in our model estimates (Figure 1); R_0_ (basic reproduction number) among studied states ranged 1.24-1.96 (95% CI: 1.36-1.78).

In addition, effectiveness of lockdown was revealed with improvement of SI (pre-lockdown: mean 3.82 days, 95% CI: 2.55-5.1; post-lockdown: mean 14.26 days, 95% CI: 2.44-26.08), DGR (pre-lockdown: mean 0.22, 95% CI: 0.11-0.34; post-lockdown: mean 0.13, 95% CI: 0.10-0.16), whereas CFR increased slightly (pre-lockdown: mean 2.54, 95% CI: 0-5.52; post-lockdown: mean 3.11, 95% CI: 1.19-5.03), as depicted in Figure 2.

**Figure 1.**
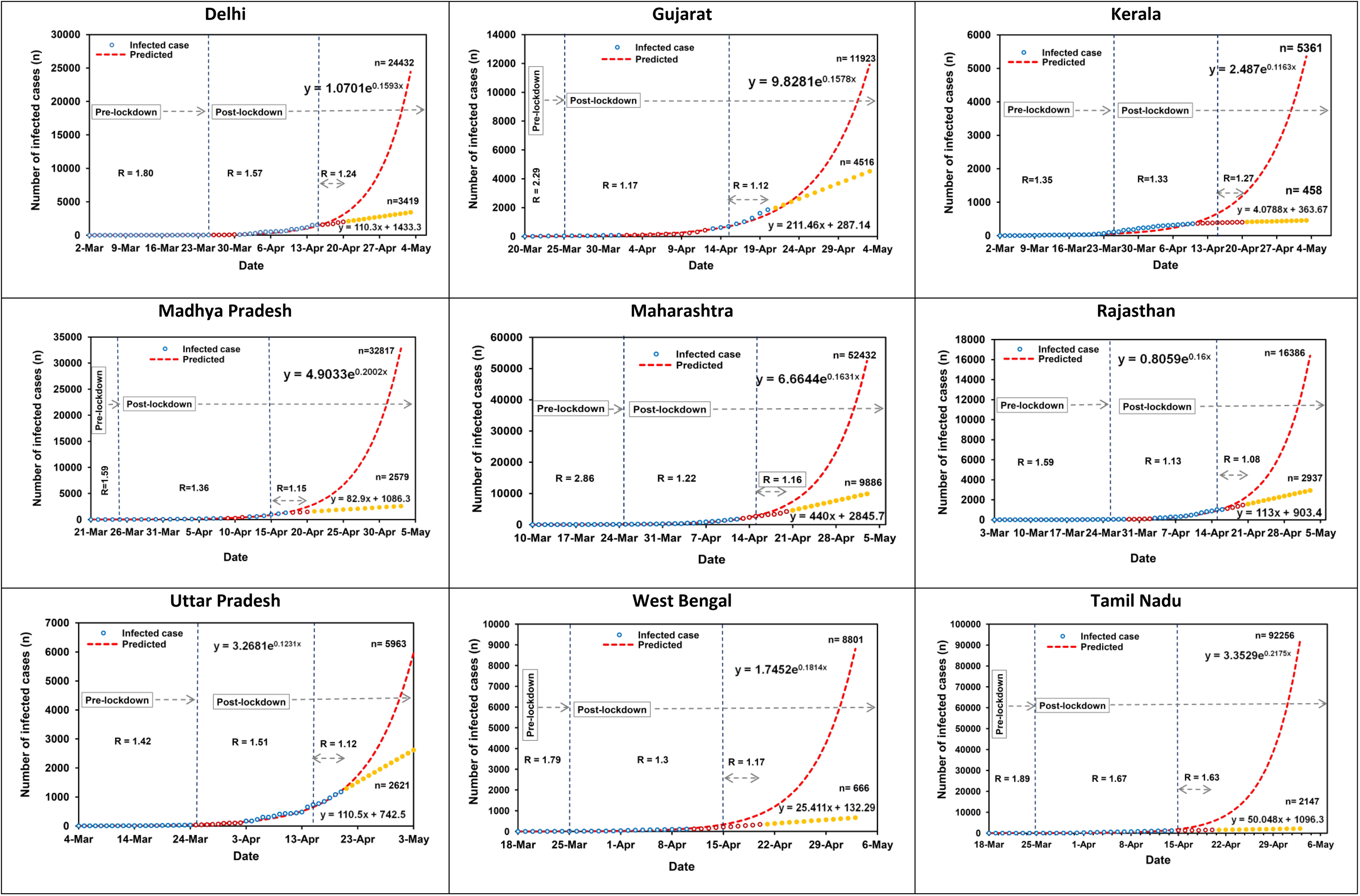
Data-based (as on April 20, 2020) and model estimation (up to May 3, 2020) of infected cases in Indian states. Abbreviation: R, Reproduction number. Red dashed lines denote estimated case numbers without lockdown, using exponential model. Circles represent observed infected case numbers with lockdown, forecast beyond April 20, 2020 (yellow solid circles) using linear model. Red hollow circles indicate lockdown-effective days, in two phases commencing on March 25, 2020 and April 15, 2020 (demarcated by dashed vertical lines)

**Figure 2.**
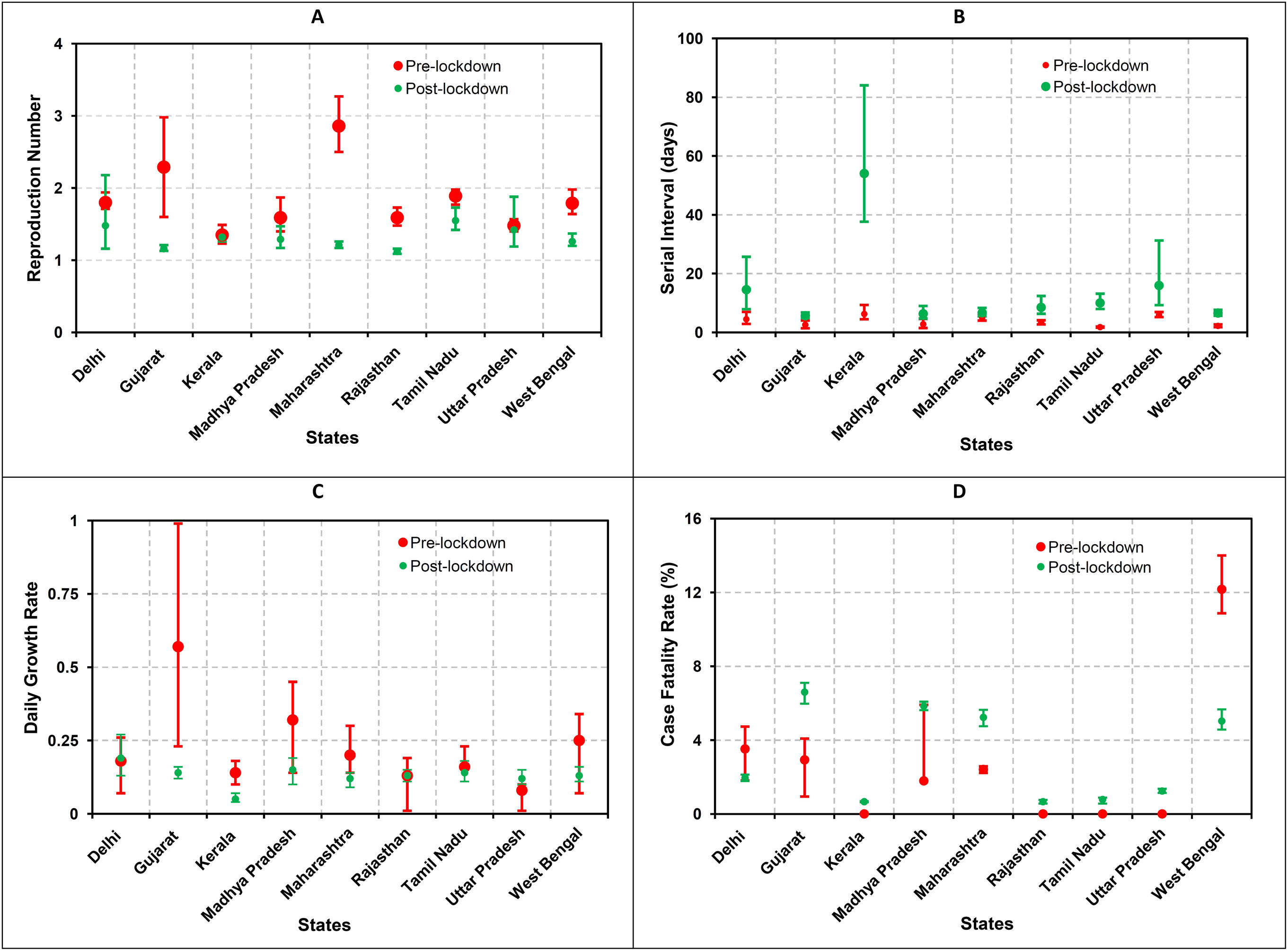
Epidemiological parameters of COVID-19 in Indian states before and after lockdown. A: Reproduction number; B: Serial interval; C: Daily growth rate; D: Case fatality rate. Vertical bars indicate standard error of mean

## Discussion

In order to monitor and foresee the plausible progress of the ongoing COVID-19 pandemic, estimation of SI, CFR and R_0_ is of prime importance,^5^ and daily deaths, and growth rate of COVID-19 cases also display importance in gaging the scale of the pandemic across time and space, and in framing policies and procedures.^6,7^

The dynamics of COVID-19 epidemic in Indian states with interventions was characterized by sub-exponential growth, which around 11 April 2020, deviated from the exponential growth of potential case numbers without interventions. Despite heterogeneities in socio-economic status and healthcare systems across Indian population,^2^ similar growth patterns of COVID-19 among the states considered in the current study imply the effectiveness of the outbreak interventions.

Emergence of pre-symptomatic transmission likely occurred before lockdown in the states except Kerala, Uttar Pradesh, and Maharashtra, as explained by SI values lower than or almost equal to the incubation period (~5 days), while SI values higher than that of the incubation period indicated symptomatic transmission post-lockdown. Delhi and Uttar Pradesh displayed reduced post-lockdown DGR, while CFR increased in the states except Delhi and West Bengal. Maharashtra, Gujarat, and West Bengal curbed the disease spread more effectively compared to the other states with inflected post-lockdown R values, while West Bengal, Madhya Pradesh, Kerala, and Tamil Nadu projected greatly reduced case estimates (92%-98%) with lockdown, as on 3 May 2020, indicating better containment. The effect of COVID-19 containment was more prominent in the second phase of lockdown with declining R values, which was still >1, suggesting further requirement of application of non-pharmaceutical interventions, in order to limit the SARS-CoV-2 transmission.^8^

## Conclusion

The sustained lockdown measures, combined with testing-isolating-contact tracing, face mask using and hygiene-practice are need of the time for an early end of the ongoing COVID-19 pandemic in Indian context. However, as the outbreaks develop further availability of information will bring a clear reflection afresh.

## Data Availability

Data utilized in the study were retrieved from the publicly accessible website of the Ministry of Health and Family Welfare, India

